# Alcov: Estimating Variant of Concern Abundance from SARS-CoV-2 Wastewater Sequencing Data

**DOI:** 10.1101/2021.06.03.21258306

**Authors:** Isaac Ellmen, Michael D.J. Lynch, Delaney Nash, Jiujun Cheng, Jozef I. Nissimov, Trevor C. Charles

**Affiliations:** Department of Biology, University of Waterloo, Waterloo, ON, Canada; Metagenom Bio Life Science Inc., Waterloo, ON, Canada

## Abstract

Detection of SARS-CoV-2 in wastewater is an important strategy for community level surveillance. Variants of concern (VOCs) can be detected in the wastewater samples using next generation sequencing, however it can be challenging to determine the relative abundance of different VOCs since the reads cannot be assembled into complete genomes. Here, we present Alcov (abundance learning of SARS-CoV-2 variants), a tool that uses mutation frequencies in SARS-CoV-2 sequencing data to predict the distribution of VOC lineages in the sample. We used Alcov to predict the distributions of lineages from three wastewater samples which agreed well with clinical data. By predicting not just which VOCs are present, but their relative abundances in the population, Alcov extracts a more complete snapshot of the variants which are circulating in a community.

## Introduction

The COVID-19 pandemic caused by the SARS-CoV-2 virus is responsible for millions of deaths worldwide. Monitoring the composition of SARS-CoV-2 lineages which are circulating in a region is crucial to managing the pandemic. Detecting variants of concern (VOCs) can help inform epidemiological models, as some VOCs have increased transmissibility and immune escape [1]. A useful approach in tracking these variants is monitoring SARS-CoV-2 in wastewater. Since viral particles are shed in stool, collecting and analyzing wastewater from wastewater treatment plants provides a snapshot of the viral composition for an entire community [2].

A sample of SARS-CoV-2 can be tested for VOCs using next generation sequencing. Clinical samples may be sequenced and aligned to a reference genome to determine which mutations are present. These signature mutations can be used to identify which lineage, and so VOC, a sample contains. However, sequencing wastewater samples is more complicated for two reasons. First, the samples usually contain multiple lineages (and possibly multiple VOCs) and so the consensus sequence can not provide an accurate snapshot of the composition of the sample. Second, wastewater samples often contain low-quality RNA which may lead to significant gaps in the sequenced genome of the virus. Thus, only relying on a small number of unique signature mutations may be an unreliable method for identifying variants of concern if these mutations are not covered in the sample.

Here we present Alcov (abundance learning of SARS-CoV-2 variants), a tool for calling VOCs in a mixed sample and estimating their relative abundance within that sample using a large number of mutations distributed across the SARS-CoV-2 genome. This allows for the estimation of VOC abundance within a community using only sequenced wastewater instead of a massive sequencing program of infected individuals.

## Methods

### Preprocessing the reads

Alcov receives a BAM file of filtered reads, aligned to the SARS-CoV-2 reference genome, NC_045512.2 [3]. In all of our experiments we preprocessed the reads using SeqPrep [4] to quality filter and merge paired reads if required. Next we aligned the reads to the reference genome using BWA [5] and used samtools [6] to sort the reads and save the file as a BAM.

### Converting amino acid mutations to nucleotide polymorphisms

Since many repositories of common SARS-CoV-2 mutations such as covariants (https://covariants.org/) and outbreak.info [7] (https://outbreak.info/) refer to mutations by their amino acid change on the gene, rather than nucleotide change on the genome, we first implemented a function to convert between the two. Briefly, to convert from a nucleotide to an amino acid change, the program finds the gene and codon for that location and returns the wildtype and mutated amino acids at that location. To convert from an amino acid change to a nucleotide change, the tool finds the amino acid and determines which single nucleotide polymorphism could account for it.

### Determining mutation frequencies

To determine the frequency of a given mutation, Alcov scans the reads at each index using pysam [8] and calculates the frequency as the number of single-nucleotide variants over the number of reads which cover that base. In order to reduce the variance of the estimates, Alcov only considers locations with coverage of at least 40 reads. There are however cases where there are multiple nucleotide polymorphisms that can account for the same amino acid change, such as alternate codons or codon deletions. In such cases, Alcov assigns the mutation’s prevalence as the highest nucleotide polymorphism frequency from the set.

### Lineage abundance estimation as an optimization problem

After calculating the frequency of each mutation, the problem is to determine a relative abundance for each VOC lineage that could collectively account for the observed mutation frequencies. Let *y*_*i*_ denote the frequency of mutation *i* and *x*_*i,j*_ denote whether lineage *j* contains mutation *i*. Finally, let β_*j*_ denote the predicted relative abundance of lineage *j* in the sample. Then the predicted frequency of mutation *i* is:

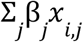

We can cast the problem of determining the lineages as determining a set of β_*j*_ to minimize the total difference between the observed mutation rates and the predicted mutation rates:

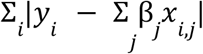

Or, similarly, the squares of the differences:

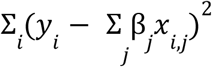

This formulation is a well studied problem in statistics known as ordinary least squares (OLS) which can be solved efficiently in closed form [9]. We implemented the model using scikit-learn version 0.24 [10] which allows for the additional constraint that all β_*j*_ must be positive. While there is no explicit condition that Σ_*j*_β_*j*_ ≤ 1, in most cases the best fit will be a reasonable estimation of abundances.

It is worth noting that this formulation addresses both of the major problems with wastewater sequencing. First, it seeks to solve the problem of determining the composition of lineages in the sample and so explicitly assumes that the reads are coming from a mixed sample. Second, it is resilient to dropout of mutations (e.g., due to sample degradation or unequal coverage). By considering each mutation as a data point in the linear regression, it can easily consider only and all mutations with sufficient read coverage.

## Results

### Data collection

In order to test the predictions made by Alcov, we searched for “SARS-CoV-2 wastewater” on the Sequencing Read Archive (SRA) (https://www.ncbi.nlm.nih.gov/sra). Seven SRA studies were found, of which three had sufficient data to make predictions. One recent sample from each study was downloaded and preprocessed using the steps described above before being passed into Alcov for VOC abundance prediction. The specific samples used were SAMN18310570, SAMN18378816, and SAMN18915228. These samples were each analyzed as a case study. The VOC mutation information Alcov uses was downloaded from https://outbreak.info/compare-lineages on May 12, 2021.

### Frankfurt, Germany - December 2020 (SAMN18310570)

The first sample analyzed was collected from Frankfurt in December 2020 [11]. This sample acted as a negative control since there were not widespread VOCs in Germany at that time. As expected, Alcov predicts negligible abundances for all of the variants of concern (Figure 1). The slightly non-zero values are likely due to sequencing/reverse transcription errors, or low levels of sequence variation within that community.

**Figure 1.**
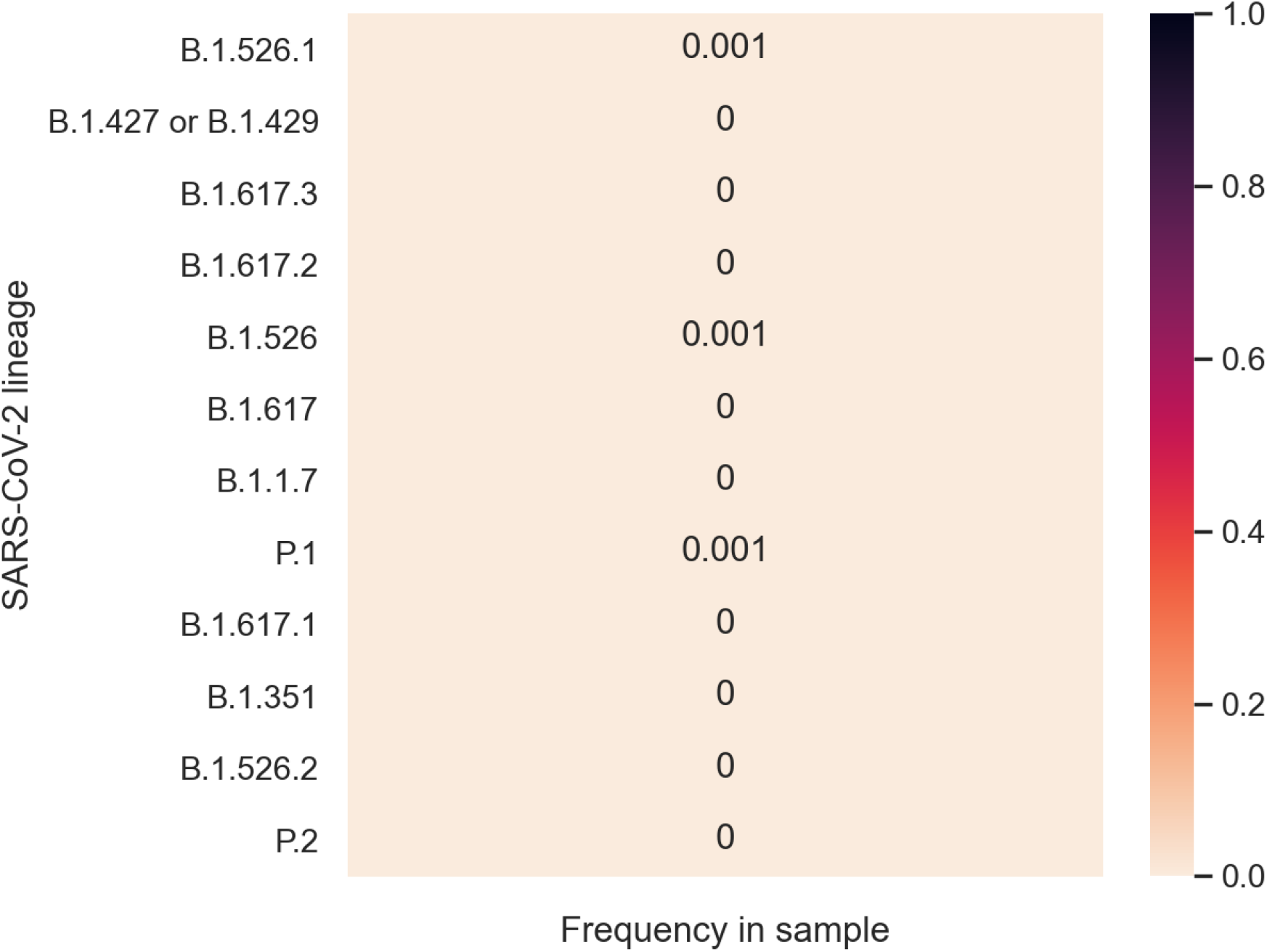
Predicted lineages in Frankfurt (December 2020). Lineages are coloured according to their predicted frequency which is also labeled. As expected, the frequency of each VOC lineage is negligible.

### New York City, New York, USA - March 2021 (SAMN18378816)

The second sample analyzed was collected in New York City (NYC) in March 2021 [12]. Only a 332-nucleotide region of the receptor binding domain was sequenced which covered six known mutations present in VOCs (S:L452R, S:S477N, S:T478K, S:E484K/Q, and S:N501Y). Because multiple VOCs were identical with respect to these mutations, some were merged before making the prediction. Two VOCs were predicted by Alcov in this sample, both of which are consistent with clinical data [7]. B.1.1.7 was predicted to account for 43% of the sample whereas some mixture of B.1.526.1, B.1.427, and B.1.429 was predicted to account for 22% of the sample (Figure 2). It is very likely that the latter portion was due to B.1.526.1, which is a strain that originated in NYC and has been observed in high prevalence in NYC clinical samples [7]. Similarly, the B.1.1.7 proportion is in line with clinical sampling data.

**Figure 2.**
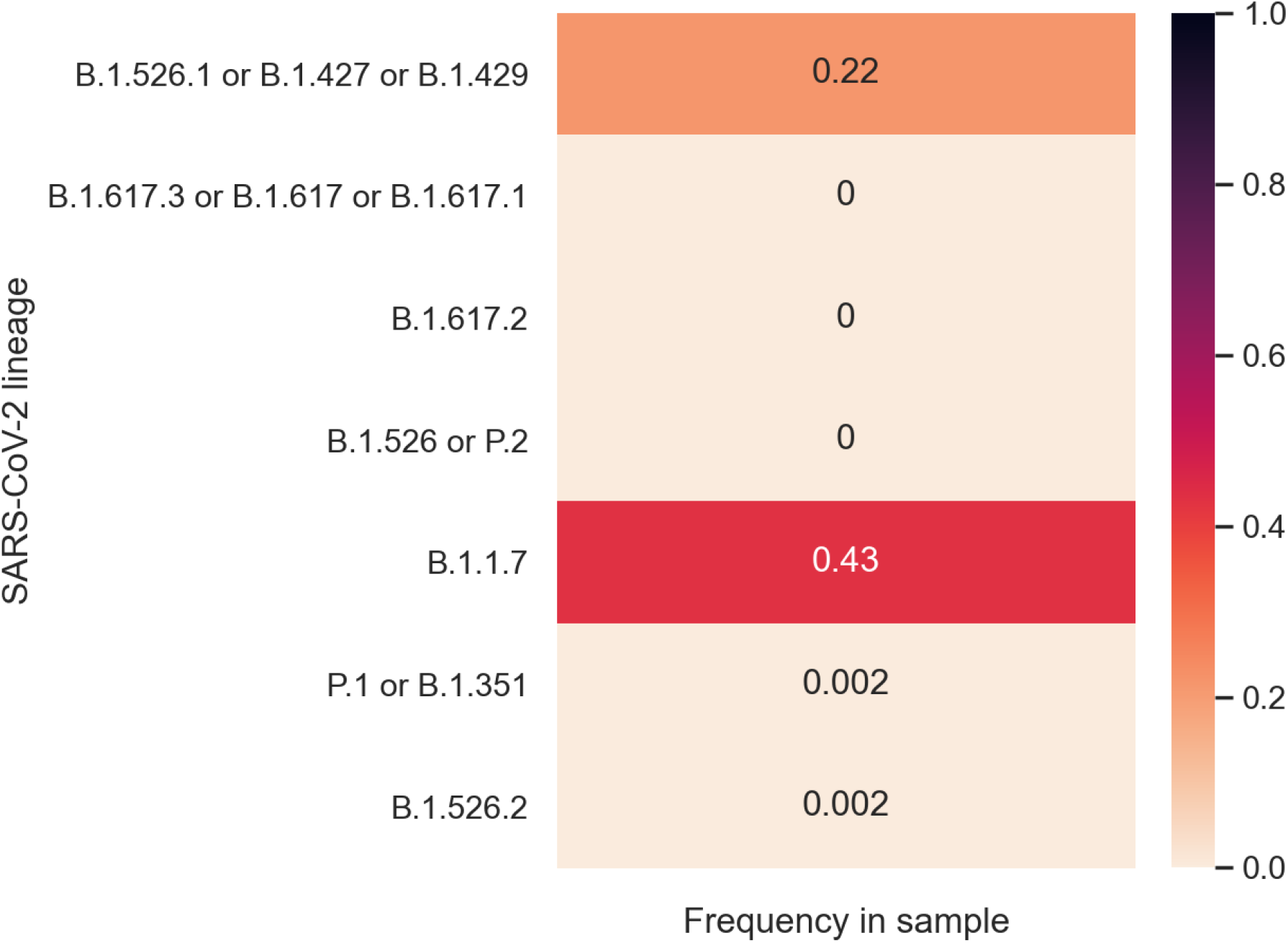
Predicted lineages in NYC (March 2021). Because of limited sequencing coverage, some VOCs were indistinguishable and so automatically merged. The B.1.1.7 and B.1.526.1 predictions agree with clinical data.

### Newport, Oregon, USA - March 2021 (SAMN18915228)

The third sample analyzed was collected in Newport in March 2021. The sample had many regions with low coverage which made it challenging to analyze by looking at only the most common characteristic mutations. [tool name] predicted a split between 62% B.1.417/B.1.429 and 38% B.1.351 (Figure 3). The B.1.417/B.1.429 prediction makes sense since these VOCs originate from California which is adjacent to Oregon and are present in clinical samples from Lincoln county which includes Newport [7]. The B.1.351 prediction is surprising since it is rare in Oregon and, to our knowledge, has not been discovered in Lincoln. Mutations which were not included in the prediction due to insufficient read depth were analyzed manually, and supported the B.1.351 prediction. This example may demonstrate some of the utility of Alcov for wastewater identifying VOCs that have yet to be captured in clinical sequencing. That said, it is unlikely that the proportions in this sample are representative of the larger community, and may have been exaggerated due to stochasticity in PCR amplification.

**Figure 3.**
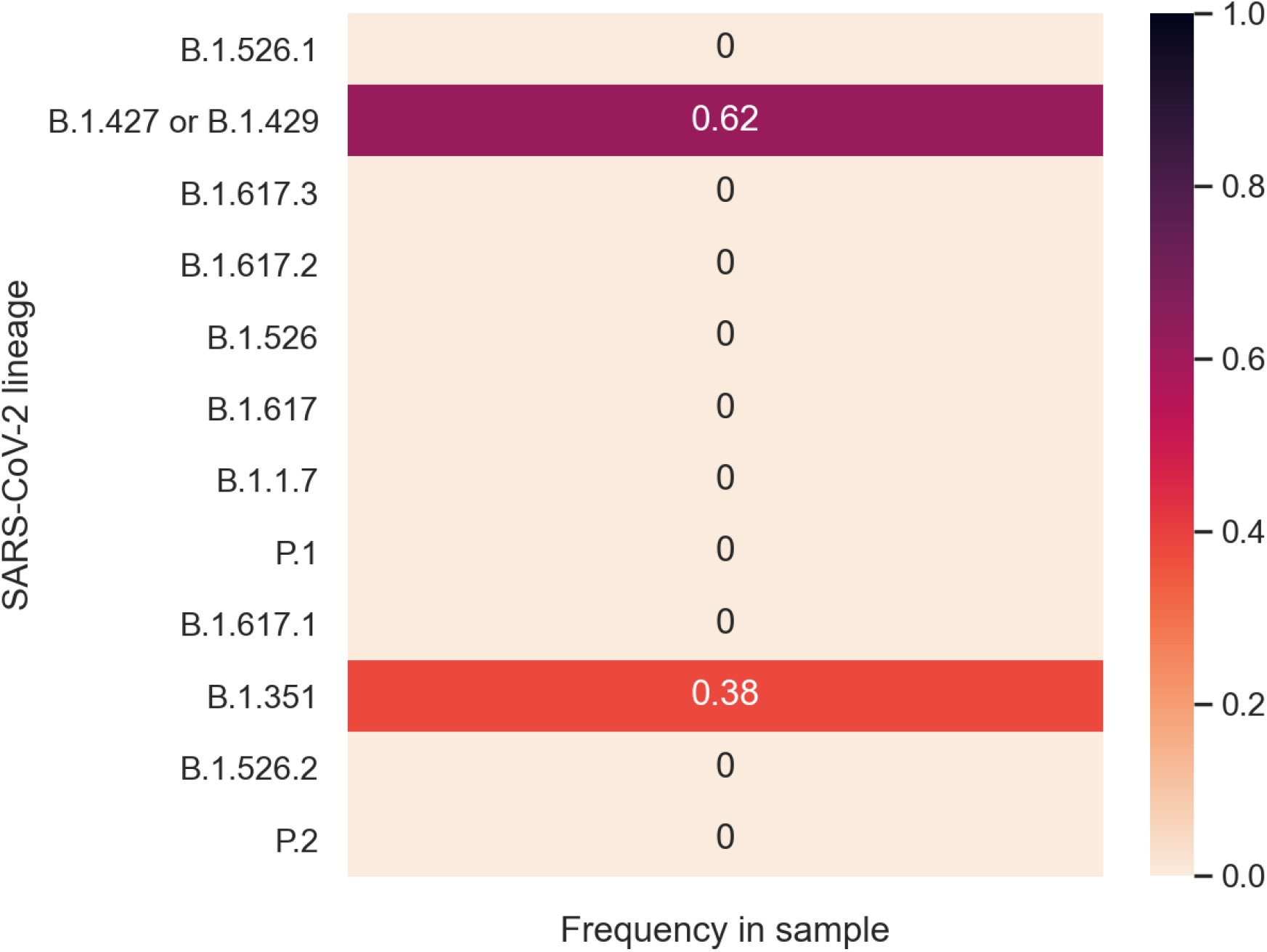
Predicted lineages in Newport (March 2021). The B.1.427/9 lineages originated in California which is adjacent to Oregon. The B.1.351 prediction is high compared to clinical data but is well supported across multiple mutations in the sample.

## Discussion

By estimating the abundance of VOCs from single wastewater sequencing runs, Alcov enables genomic epidemiology of SARS-CoV-2 variants without the need for a large number of clinical sequences. Thus, sequencing and analyzing wastewater samples is a cost-effective and rapid solution for determining the composition of variants in both large and small communities. Additionally, since clinical samples are typically only sequenced after testing positive for other diagnostics such as PCR tests, this may be a less biased population analysis. It is important to note that Alcov can only determine the composition of RNA reads which are sequenced. This composition may differ from the actual distribution of virus lineages in a population due to increased shedding, differential PCR amplification of some amplicons, differential RNA stability in wastewater, or other factors. Determining the correlation between the wastewater abundances and clinical samples will be an important avenue for future research. Another limitation is the limited lineage information which is included. Since Alcov only looks for VOCs, it may attribute the presence of mutations from non-VOC lineages to VOCs. This could be addressed by also including the specific lineages which are circulating in the community which is being sampled (as determined by clinical sequencing data). Finally, it is worth noting that the approach taken by Alcov is not specific to SARS-CoV-2. The tool could be adapted to monitor future pandemics, or even plant viruses in hydroponic systems.

## Data Availability

Data IDs which are used are provided. Source code is linked.

https://github.com/Ellmen/alcov

## Availability

Alcov is written in Python and can be used as a command line interface. It has been tested on macOS and is available for download at https://github.com/Ellmen/alcov.

## Acknowledgments

This work was supported by Mitacs Accelerate Fellowships (IT18981) to IE and DN, through funding held by JIN.

